# Optimal Selection of COVID-19 Vaccination Sites at the Municipal Level

**DOI:** 10.1101/2021.06.20.21259194

**Authors:** Kurt Izak M. Cabanilla, Erika Antonette T. Enriquez, Renier Mendoza, Victoria May P. Mendoza

## Abstract

In this work, we present an approach to determine the optimal location of coronavirus disease (COVID-19) vaccination sites at the municipal level. We assume that each municipality or town is subdivided into smaller administrative units, which we refer to as villages or barangays. The proposed method solves a minimization problem arising from a facility location problem, which is formulated based on the proximity of the vaccination sites to the villages, number of COVID-19 cases, and population densities of the villages. We present a numerical scheme to solve the optimization problem and give a detailed description of the algorithm, which is coded in Python. To make the results reproducible, the codes used in this study are uploaded to a public repository, which also contains complete instructions on how to use them. As an illustration, we apply our method in determining the optimal location of vaccination sites in San Juan, a town in the province of Batangas, in the Philippines. We hope that this study may guide the local government units in coming up with strategic plans for the COVID-19 vaccine rollout.

## 1. INTRODUCTION

The coronavirus disease (COVID-19), which was first reported in Wuhan, China, has spread across the globe and was declared a pandemic by the WHO on March 11, 2020 [1,2]. It has been shown that the COVID-19 vaccines are effective against severe disease and initial findings suggest that they can protect the population against infection [3-5]. An effective vaccination campaign can reduce the probability of disease resurgence and alleviate the economic burden of the pandemic [6]. It is estimated that 60-70% of the population must be fully vaccinated to achieve herd immunity, a threshold that provides indirect protection to uninfected individuals from getting COVID-19 from infected hosts [7-9]. To achieve the target immunity, the community’s hesitancy over the COVID-19 vaccines must be addressed by the policymakers [10]. In a survey conducted in January 2021 by the Pulse Asia, almost half of the Filipino respondents said that they are not willing to get vaccinated against COVID-19 because of safety concerns. Hence, a collaborative effort among several stakeholders and sectors is needed to address this issue [11]. Unfortunately, COVID-19 vaccination in the Philippines is progressing at a very slow rate due to the inadequacy of the policymakers’ long-term strategic plan [12]. We hope that this study may guide the Philippine local government units in coming up with strategic plans for vaccine rollout. We propose a way to select optimal vaccination sites from already existing facilities to make the vaccines more accessible to the public and achieve herd immunity at the soonest possible time.

Our proposed approach solves a facility location program, which is a problem that minimizes the cost of satisfying a set of demands with respect to some set of constraints [13]. Facility location problem has a variety of applications in determining optimal locations of solar power plant sites [14], hydrogen production sites [15], tsunami sensors [16,17], infrastructure maintenance depot [18], tower sites for early-warning wildfire detection systems [19], and high-speed train stations [20], among others. Facility location has also been used in several COVID-19-related studies. In [21], an optimal allocation of COVID-19 testing kits among accredited testing centers has been proposed. The optimal location of pharmacies for COVID-19 testing to ensure access has been studied in [22]. Identification of locations of COVID-19 emergency logistic centers has been proposed in [23]. In [24], optimal COVID-19 testing facility sites in Nigeria have been studied.

Based on our review of related literature, very few studies have been conducted on the applications of facility location problems in COVID-19 vaccination distribution strategies. In [25], an approach to optimize vaccine distribution strategies has been proposed by selecting locations that will minimize the death toll. The method relies on an epidemiological model to capture the effects of vaccinations and mortality caused by COVID-19. In [26], a mathematical framework in finding the optimal locations of distribution centers for test kits and vaccines has been developed. In [27], a linear programming model was used for COVID-19 vaccine allocation in the Philippines at the national level. Because of the scale of these studies, constraints must be considered (e.g., shipping cost, production capacity, operating cost, etc.). In this study, we consider a local-scale vaccination strategy. By doing so, we can remove the logistic constraints associated with the delivery of the vaccines. Thus, we can focus on finding the optimal location of vaccination sites that will make the vaccines more accessible to the population of the town or city. Our work can be used in conjunction with vaccine allocation methods at the national level [25-27]. Once the vaccines are allocated to a town, our method can be applied to identify the sites where the vaccines will be distributed.

In the next section, we formulate the mathematical optimization model to address the vaccination site location problem. Then, we present the numerical method that solves the minimization problem and the open-source software that we developed. The open-source software is designed to be user-friendly and easy to implement. We illustrate how our proposed method works by applying the program in identifying optimal vaccination sites in San Juan, Batangas, Philippines. Finally, we present our conclusions and recommendations for future research.

## 2. OPTIMIZATION PROBLEM

Our goal is to determine the optimal location of *L* vaccination sites in a town from a list of *M* possible vaccination sites. We consider existing facilities such as public schools and hospitals as possible vaccination sites. Furthermore, suppose that the town is divided into *N* administrative units, which we refer to as villages. These are usually the country’s basic units of government. In the Philippines, all towns are composed of several administrative units called barangays. Let {*V*_*i*_ : *i* = 1,2, …, *M*} be the set containing the locations of the possible *M* vaccination sites. Each *V*_*i*_ is represented by a two-dimensional vector whose components are the latitude and longitude of the *i*th vaccination site. Define {*B*_*j*_ : *j* = 1,2, …. *N*} as the set containing the location of the *N* administrative units. We can set *B*_*j*_ as the location of the village/barangay hall or the community center, which is usually situated at the center of the village. Similarly, each *B*_*j*_ is a two-dimensional vector whose components are the latitude and longitude of the *j*th village or barangay.

Define *d*(*V*_*i*_, *B*_*j*_) as the distance of the vaccination site *V*_*i*_ from the barangay hall *B*_*j*_. In facility allocation problems, different distance measures are used. For example, the Euclidean distance was used in [28]. It was argued in [29] that the *l*_1_ distance (also known as Manhattan distance) more accurately models the driving distance in a city road network. However, in rural towns, the roads may not follow a rectangular grid pattern. Since *V*_*i*_ and *B*_*j*_ are accessible via the road network of a town, we utilize a Python package to calculate the actual driving distance from *V*_*i*_ to *B*_*j*_. This approach makes the computation of distance more realistic.

Now, suppose *L* = 1, that is, only one vaccination site is assigned to the whole town. Then, one distribution strategy is to choose the vaccination site that lies the closest to all the *B*_*j*_ ′*s*. That is, we solve

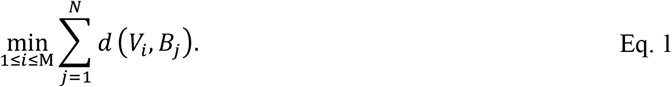

However, the minimization problem in Eq. (1) does not take into consideration the population of the villages. To resolve this, we add more weight on the vaccination sites that are closer to the more populous areas of the town.

Define *P*_*j*_ as the population of the *j*th village and *T*_*p*_ as the total population of the town. Then 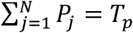. Hence, we solve

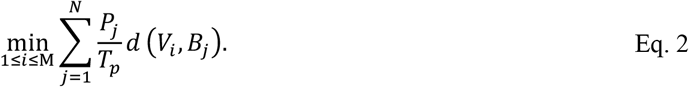

Moreover, we want to place the vaccination sites near villages with high numbers of confirmed COVID-19 cases. Define *C*_*j*_ as the number of confirmed COVID-19 cases in the *j*th village and *T*_*c*_ as the total number of confirmed COVID-19 cases in the town. Then 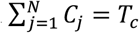. Similar to how population is incorporated in Eq. (2), we add weights on the villages with high number of confirmed COVID-19 cases. Thus, we solve

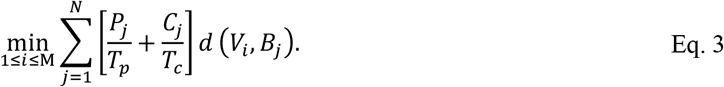

Next, we consider the case when the number of vaccination sites is more than 1, that is, *L* ≥ 2. If there are *L* vaccination sites, we want the resident of the *j*th village to go to the nearest vaccination site. We can generalize the minimization problem in Eq. (3) as follows:

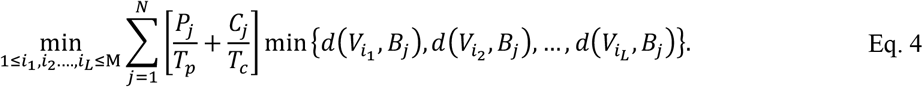

In the next section, we discuss how the minimization problem above is solved numerically.

## 3. NUMERICAL METHOD

For the overall numerical computation and some of the data extraction, we utilized the ease of use and availability of advanced open-source packages of the Python programming language. To compute the driving or road distance between two points, we leverage Open Street Maps (OSM) and its corresponding Python package OSMNX. OSM is a dynamic repository of detailed map data such as road level data, buildings, and even natural geographic objects such as rivers and mountains. OSM is built and continues to be actively updated by contributors from diverse backgrounds such as hobbyist mappers, disaster risk experts, and GIS professionals. OSM is open source, which means anyone can access and use the full breadth of its data. OSMX uses OSM data in conjunction with network graphs for a wide range of applications, such as all kinds of urban traffic and planning, all in a network graph analysis framework.

To solve the optimization problem for a given town, the user must input two files: the *village centers table* and the *vaccination centers table*. The village centers table contains the number of COVID-19 cases, population, and location of all the villages in the town. It is a file with the schema given in Table 1. The vaccination centers table contains the location of the all the possible vaccination sites. This is a CSV file with the schema shown in Table 2.

**Table 1.** The village centers table contains the number of COVID-19 cases, population, latitude, longitude, and names of all the villages or barangays in a town.

| Infected<br>(data type: integer) | Population<br>(data type: integer) | Latitude<br>(data type: float) | Longitude<br>(data type: float) | Barangay_name<br>(data type: string) |
| --- | --- | --- | --- | --- |
| Number of COVID-19 cases in the village/barangay | Population of the village/barangay | Latitude of the village hall/ community | Longitude the village hall/ community | Name of the village/barangay |
|  |  | center/ barangay<br>hall | center/ barangay<br>hall |  |

**Table 2.**
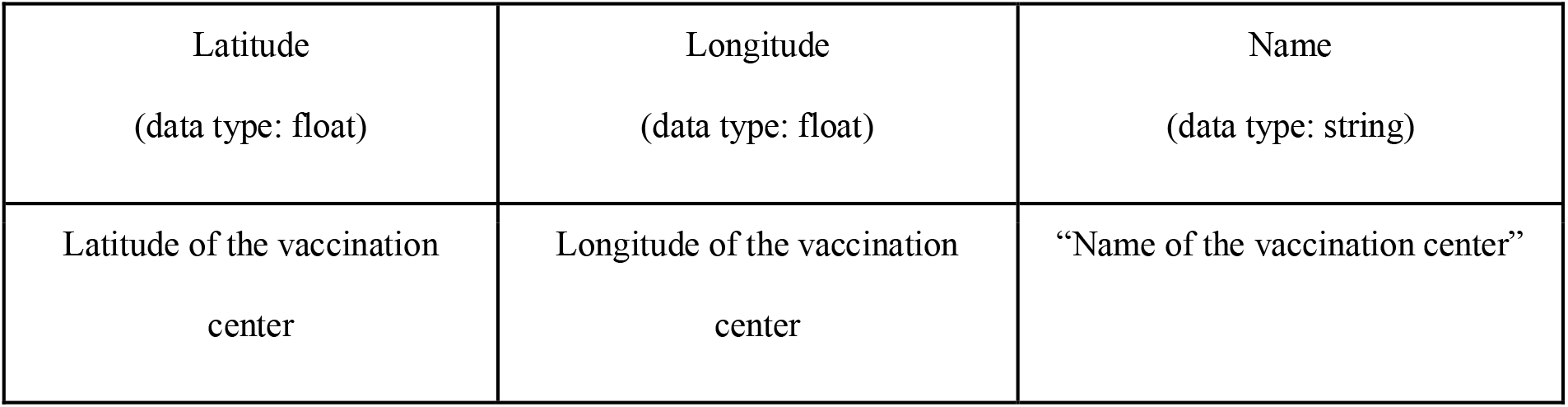
The vaccination centers table contains the latitude, longitude, and names of all possible vaccination sites in a town.

| Latitude<br>(data type: float) | Longitude<br>(data type: float) | Name<br>(data type: string) |
| --- | --- | --- |
| Latitude of the vaccination<br>center | Longitude of the vaccination<br>center | “Name of the vaccination center” |

We found that it is possible to automate the extraction of the latitude and longitude data for the vaccination centers table using OSM to a considerable extent. However, the OSM automation could not differentiate between public and private schools, thus necessitating some manual review. OSM automation can be used to generate an initial version of the vaccination centers table on which the end-users can then build on by adding or removing vaccination centers to be considered. Even though the automation is only partial, it will still significantly reduce the manual processing needed to obtain a sufficiently good vaccination centers table. On the other hand, OSM could not identify the village halls, so manual extraction of this data using Google Maps is needed. Careful extraction must be done, e.g., via Google Street View, as there are some discrepancies in Google Maps on the location of the village halls.

The cost function is computed directly as shown in Eq. (4), where the road distance *d*(*V*_*i*_, *B*_*j*_) between the *i*th vaccination site and the *j*th village hall, is computed via OSMX in Python. For both the single center and the *n*-site optimization, we iterate through every possible combination of all the vaccination sites and villages so that the resulting optimum is the global optimum.

The Python program we developed takes in the two tables previously mentioned and outputs a ranked list of the vaccination centers and their respective costs. Since it is already ordered by cost, the optimum would be the first row.

The code and a tutorial for the implementation of the numerical optimization method are found in the following GitHub repository: https://github.com/kurtizak/Covid-Site-Optimization. Sample xlsx files of the inputs can also be downloaded from this repository. The users can simply modify the Microsoft Excel files for easier implementation.

## 4. RESULTS AND DISCUSSION

To illustrate how our proposed method works, we find the optimal placement of vaccination centers in San Juan, a town in the province of Batangas, Philippines. San Juan is comprised of 42 barangays. A map detailing the location of the barangays in San Juan is shown in the supplementary file. Hence, {*B*_*j*_, *j* = 1,2, …, 42} contains the locations of the 42 barangay halls in San Juan. The latitudes and longitudes of the barangay town halls were manually obtained from local government directories and Google maps. Hospitals in San Juan are listed as possible vaccination sites. Since face-to-face classes are still suspended in the Philippines, public schools (elementary, high school, and college) are also listed as possible vaccination sites [30]. In January 2021, the Catholic Bishops’ Conference of the Philippines offered to transform churches in the country as COVID-19 vaccination sites [31]. The data on the latitudes and longitudes of the hospitals, schools, and churches are obtained from the Philippine Department of Health and Department of Education directories, and Google maps. A total of 65 sites were identified in San Juan, consisting of 5 hospitals, 42 elementary schools, 13 junior high schools, 2 senior high schools, 2 universities, and 1 church. Hence, {*V*_*i*_, *i* = 1,2, …, 65} contains the location of all the 65 possible vaccination sites.

San Juan, Batangas has a projected population of 125,252 in 2021 [32]. Meanwhile, as of 31 May 2021, San Juan recorded a total number of 579 confirmed COVID-19 cases. The complete information on the locations of possible vaccination sites and barangay halls in San Juan, the number of COVID-19 confirmed cases per barangay, and the population of San Juan per barangay can be found in the Supplementary file.

For a sample implementation, we consider selecting 1 to 4 vaccination sites among {*V*_*i*_, *i* = 1,2, …, 65}, that is, *L* = 1, 2, 3, *or* 4. Figures 1 and 2 show samples of the village centers table and the vaccination centers table, respectively, which are the required inputs from the user.

**Figure 1.**
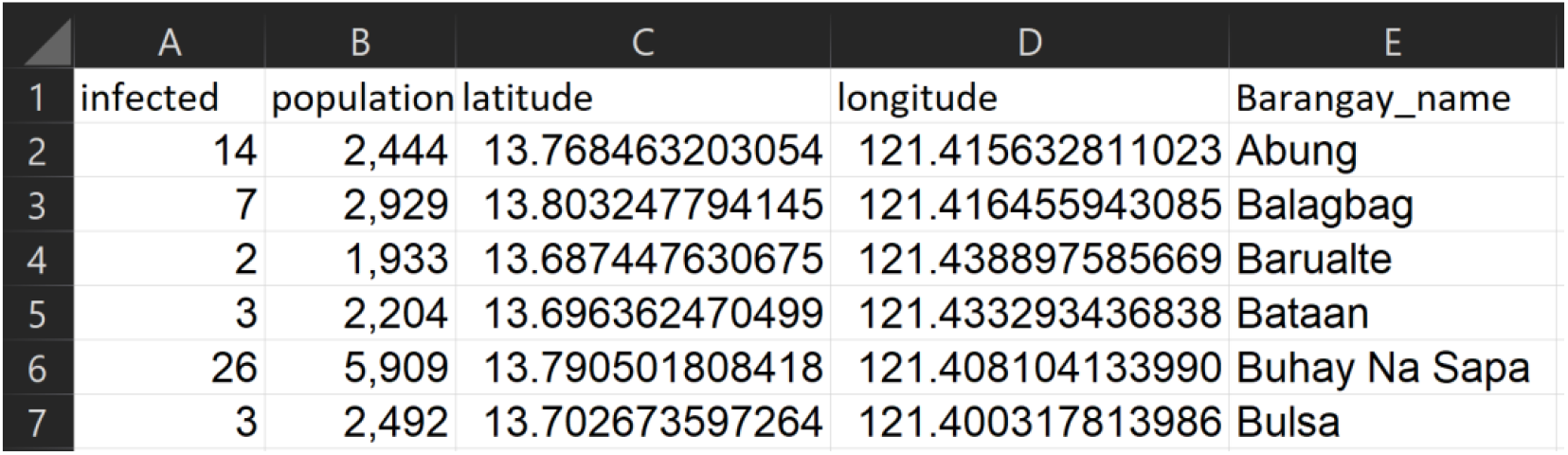
A sample Microsoft Excel file containing the data on the number of COVID-19 cases, population, location, and names of six villages or barangays in San Juan, Batangas, Philippines. This is the first of two inputs required by the system.

**Figure 2.**
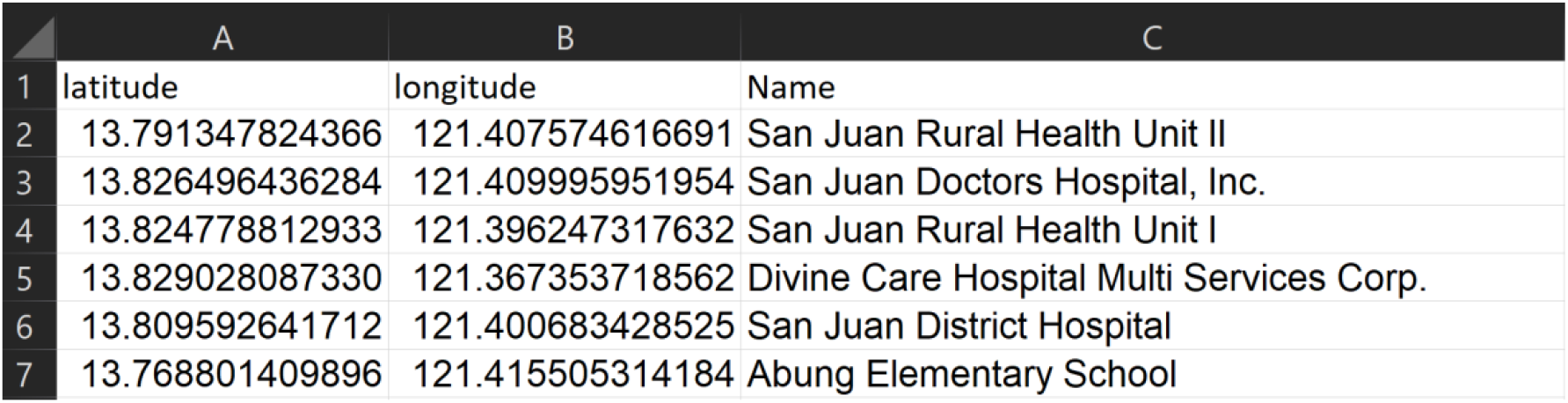
A sample Microsoft Excel file containing the data on the location and names of some of the possible vaccination sites (hospitals and public schools) in San Juan, Batangas, Philippines. This is the second of two inputs required by the system.

The code has two outputs. First, the geographic map of the area with the locations of the vaccination sites and barangay halls. Second, a data frame showing the vaccination site assignments of each barangay, as well as the distance between them. These results can be easily exported as a csv, excel, or any other format the user prefers.

Figures 3 and 4 show the geographic distribution of the optimal vaccination sites in San Juan, Batangas along with their corresponding assigned barangays for *L* = 1 and 2 sites, and *L* = 3 and 4 sites, respectively. The stars represent the optimal vaccination sites while the circular nodes are the barangay halls. All barangays assigned to a particular vaccination site have the same color. We observed that for *L* = 1, the optimal site location is close to the most populous area, which is in the northern part of the town. For *L* = 2, one optimal site is in the north (yellow star) and the other optimal site is in the south (purple star). The barangays assigned to the vaccination site in the north are represented by yellow dots, while the barangays assigned to the vaccination site in the south are represented by purple dots. As expected, the vaccination sites become more spaced out as the number of sites increases. In all cases, the optimal locations obtained are all situated in the national highway. This is also expected because the problem is formulated to minimize the driving distance from the barangay halls to the centers. Notice that for *L* = 4 sites, the two vaccination sites from the three sites did not change. The site in the northern part of the town was replaced by two sites. This is expected because the northern part of the town is the most populated and has the greatest number of confirmed COVID-19 cases (see the supplementary file). Figure 5 illustrates a sample data frame output of the vaccination centers for ten barangays in San Juan assuming that there are only 2 vaccination sites. For instance, barangay ‘Abung’ is assigned to the vaccination site named ‘San Juan Rural Health Unit 1’. The distance between the barangay and the assigned vaccination center is 6692.14 meters. Similarly, barangay ‘Barualte’ is assigned to the vaccination site ‘Paaralang Elementarya ng Bataan’ and the distance between them is 2693.79 meters. Observe that the distance between barangay ‘Bataan’ and its assigned vaccination site is 0 meters. This is because the barangay hall of Barualte and the elementary school of Bataan are in the same compound.

**Figure 3.**
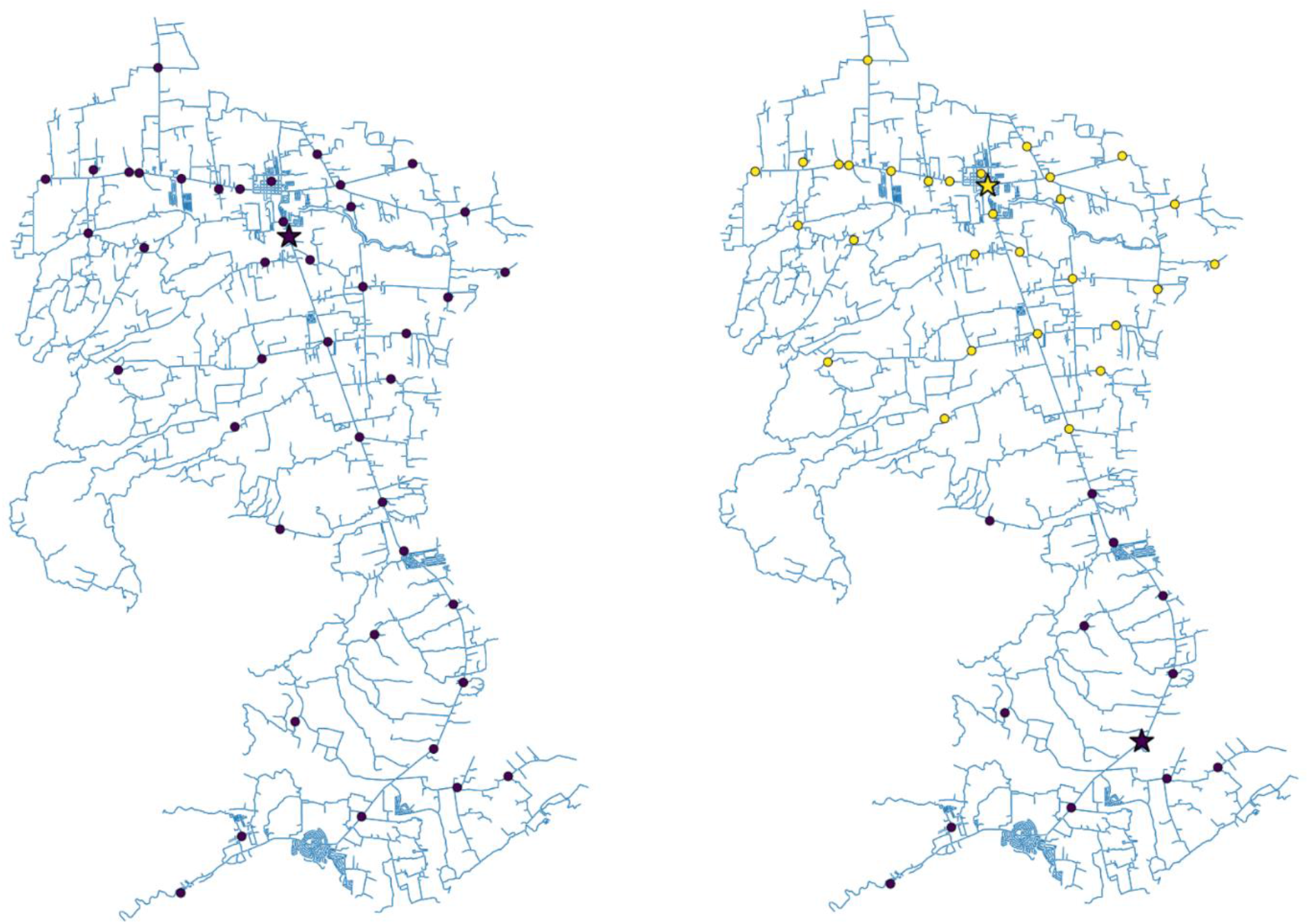
The roadmap of San Juan, Batangas showing the optimal locations of one (left) or two (right) vaccination sites. The dots represent the village/barangay halls while the stars are the computed optimal vaccination sites. The colors depict the vaccination site assignment of each village/barangay in the town.

**Figure 4.**
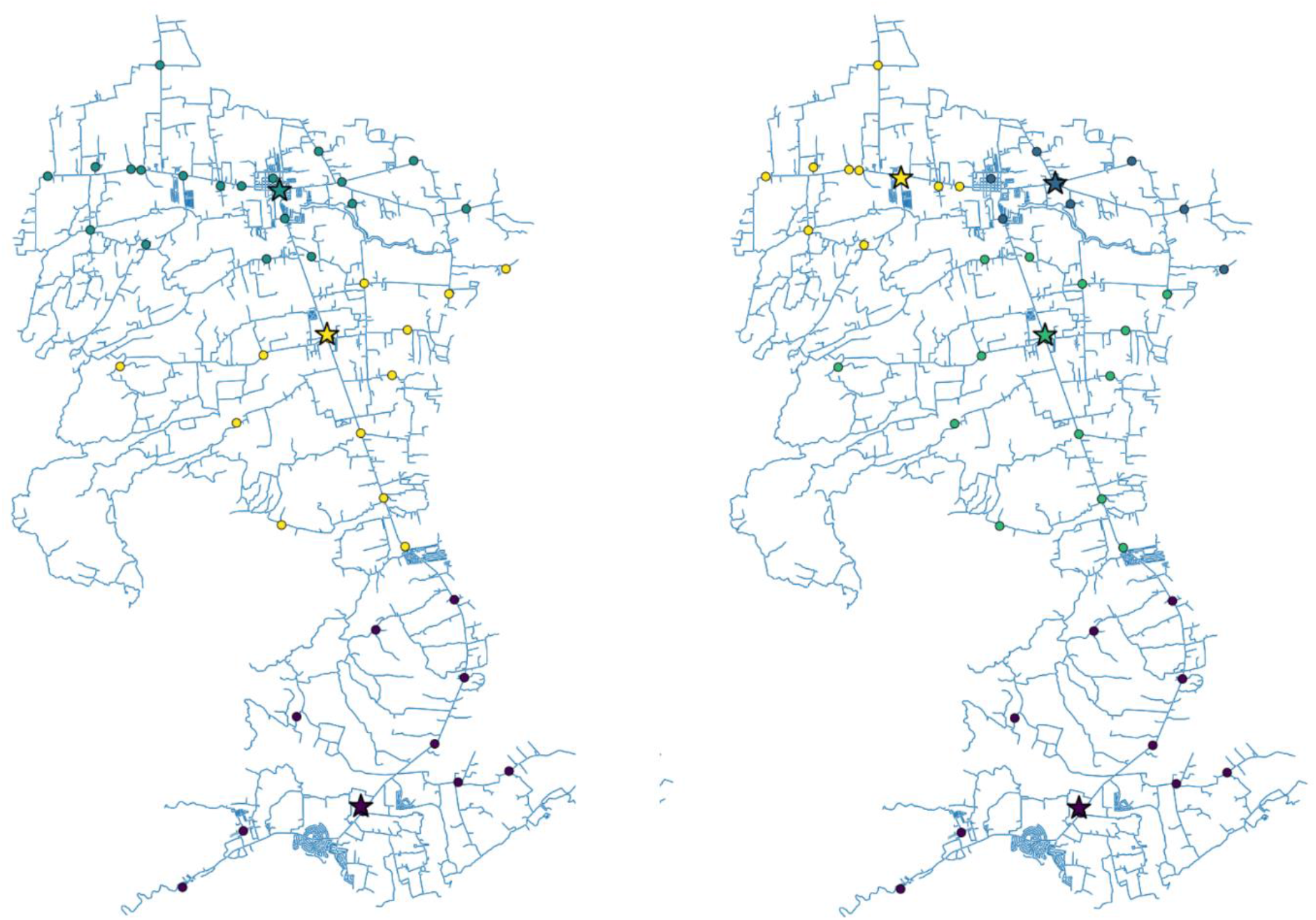
The roadmap of San Juan, Batangas showing the optimal locations of three (left) or four (right) vaccination sites. The dots represent the village/barangay halls while the stars are the computed optimal vaccination sites. The colors depict the vaccination site assignment of each village/barangay in the town.

**Figure 5.**
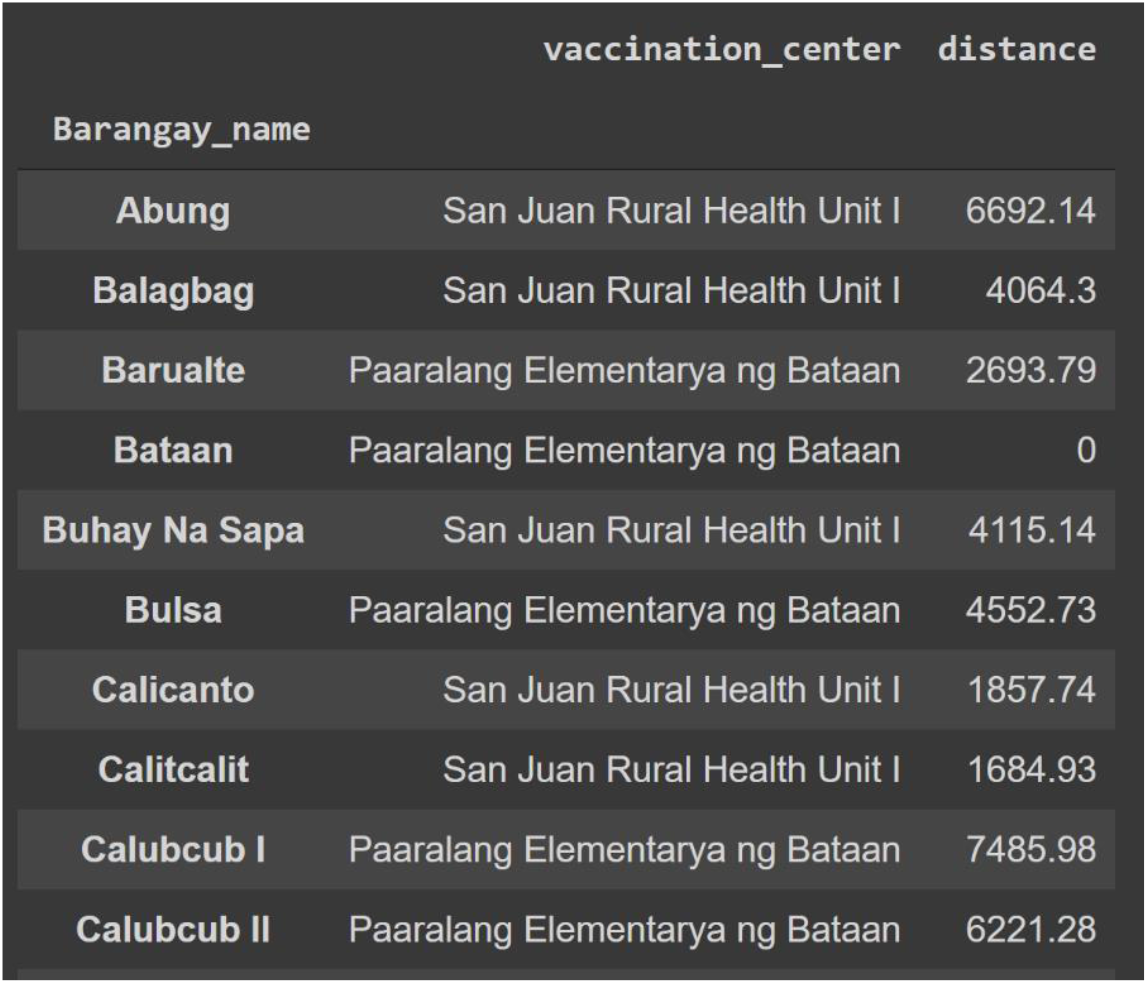
Sample output of the algorithm showing ten barangays in San Juan, Batangas and the assigned vaccination site based on proximity. The road distance (in meters) between the barangay and the assigned optimal vaccination site is also shown.

Figure 6 shows the average distance (in kilometers) of the barangays in San Juan, Batangas to the assigned optimal vaccination site, for *L* = 1, 2, 3, *or* 4 sites. On average, the difference between the road distance for one and two sites is approximately 3 kilometers while the difference between three and four sites is 600 meters. The trend shows that as more vaccination sites are opened, accessibility to the vaccines, in terms of distance, is improved. However, opening more sites has associated operational costs. Results in Figure 6 can provide information for the policymaker on finding a balance between accessibility and cost-effectiveness related to the number of vaccination sites to open.

**Figure 6.**
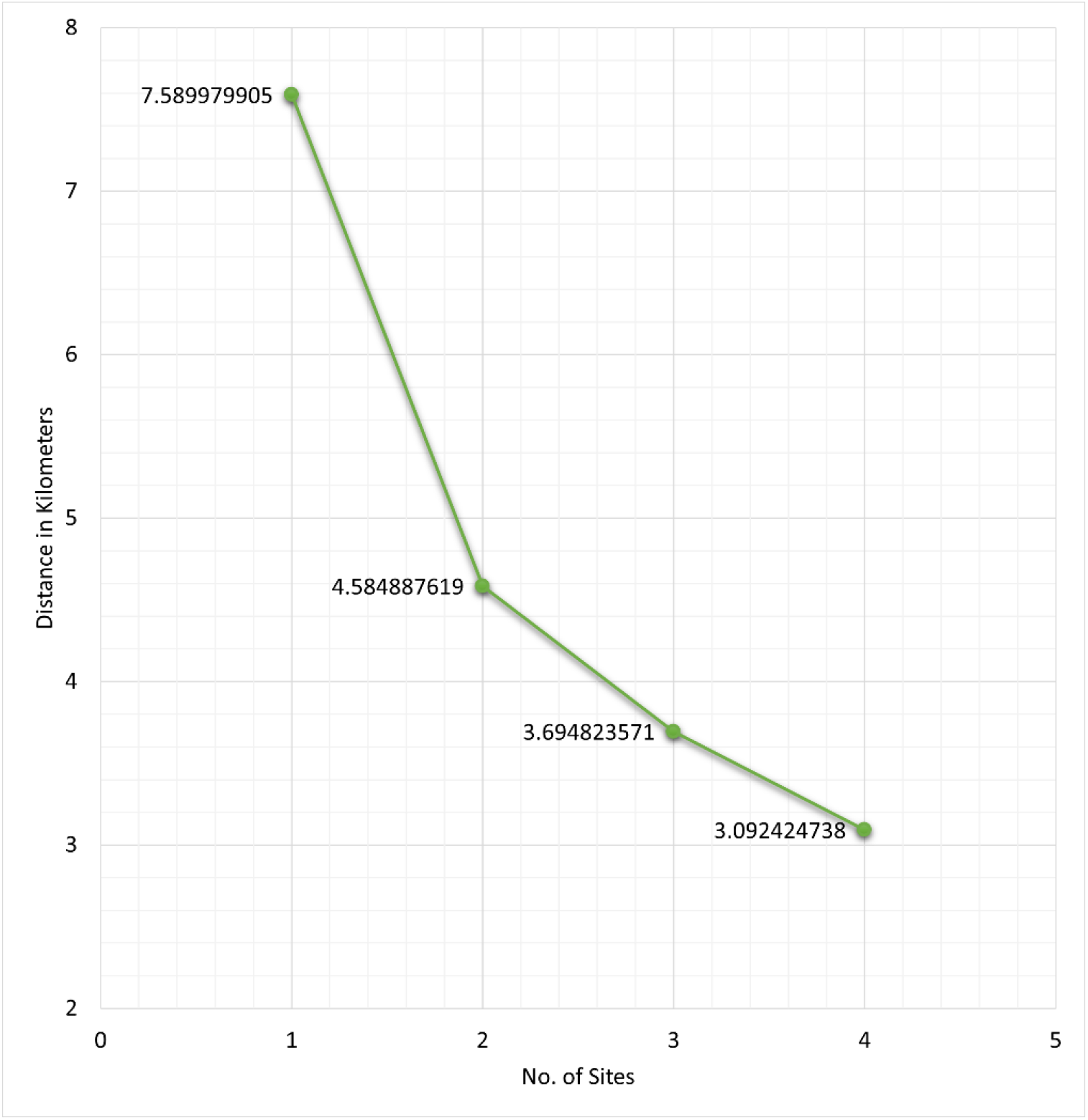
Average road distance (in kilometers) of the barangays in San Juan, Batangas to the obtained optimal vaccination site for *L* = 1, 2, 3, *or* 4 sites.

Assuming a constant vaccination rate, we can determine the number weeks it takes for a town to reach a target number of people to be vaccinated. Suppose a site in San Juan can inoculate 200 individuals per day. This rate is based on the vaccination rate of the University of the Philippines Diliman gym in Quezon City [33]. Figure 7 shows the number of weeks it takes to vaccinate 70% of the population of San Juan for *L* = 1, 2, 3, *or* 4 sites. If there is only 1 vaccination site, it takes around 62 weeks to inoculate 70% of the population in San Juan, while increasing the number of sites to 2 shortens the number of weeks to 44. Observe that the difference in time between three and four sites is only 7 weeks. If the local government has the capacity to hold vaccinations at three sites only and wishes to achieve the target of vaccinating 70% of the population in 21 weeks (same length of time as in four sites), then they can ramp up the vaccination rate at the three sites by 34.5% or by vaccinating additional 69 people per day in the three sites.

**Figure 7.**
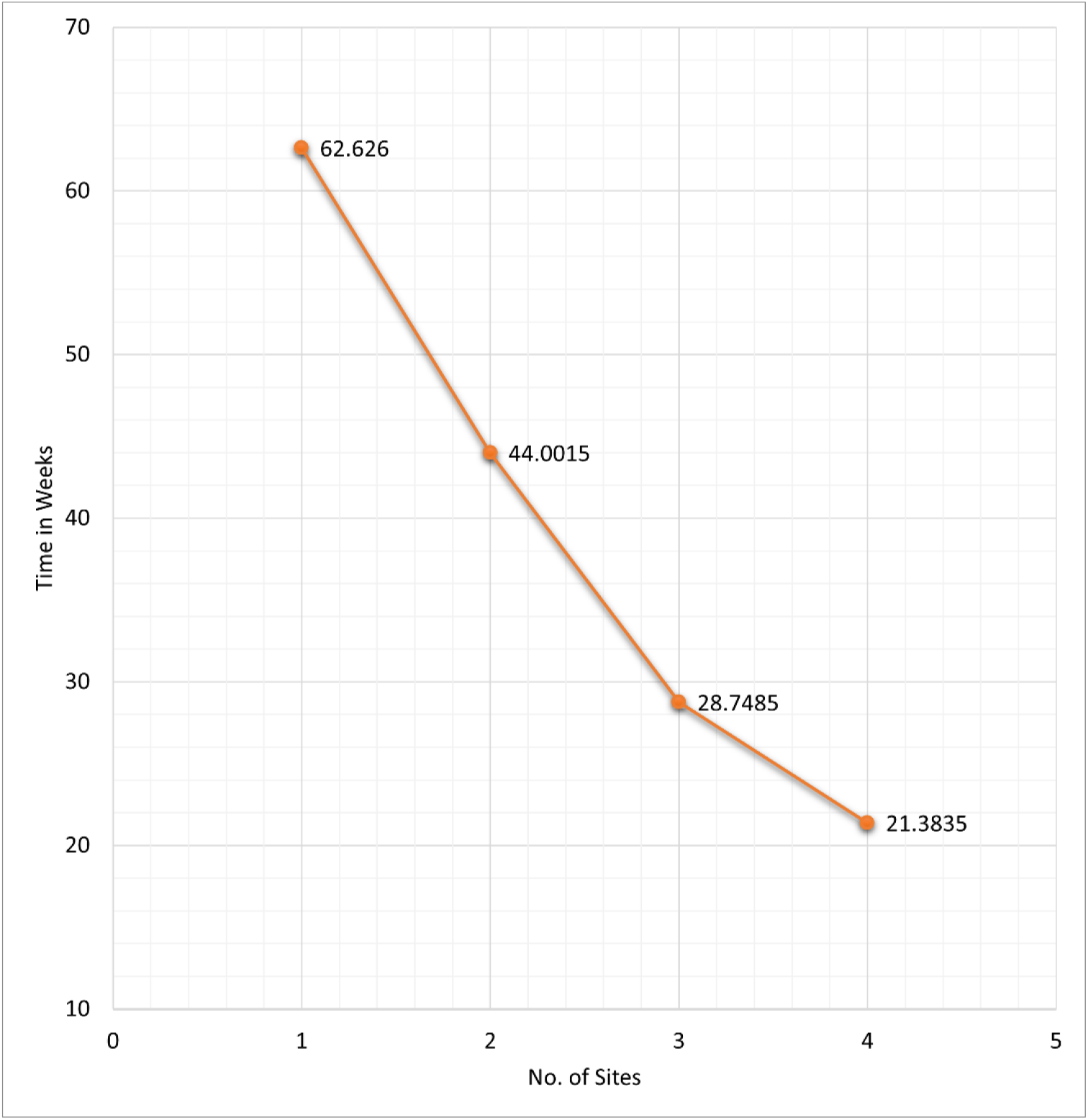
The time needed to inoculate the first dose of COVID-19 vaccines to 70% of the population of San Juan, Batangas for *L* = 1, 2, 3, *or* 4 sites.

## 5. CONCLUSION AND FUTURE WORK

Building vaccination sites uses up resources and delays the goal of achieving herd immunity. Hence, it is reasonable to set up vaccination sites from already existing facilities. In this study, we proposed an approach to strategically select COVID-19 vaccination sites at the municipal level to bring the vaccines closer to the people who need them. In finding the optimal location of the COVID-19 vaccination sites, the method considers the location of the sites, the population density of the town, and the number of COVID-19 cases per village or barangay. An open-access software has been created to make the results reproducible. The code only requires two files, one for the data on the list of possible vaccination sites and the other for the data on the villages or barangay. Our numerical simulations show strategic placements of vaccination sites to make the vaccines more accessible to the people and urge them to get vaccinated as soon as possible. The method can be beneficial to underdeveloped rural towns in developing countries, where public transportation is not reliable (or in some cases, not available).

In the illustration, we considered at most 4 vaccination sites, but the software can be easily extended to more accommodate more sites. However, this would require more computational time. Since our method is enumerative, parallel computing can be used to address the computational cost in case one wants to use our method to determine optimal locations of vaccination sites in big cities or at the provincial level. If a computing facility is not available, one can use other optimization algorithms (like meta-heuristic algorithms). Exploring other algorithms that can solve the optimization problem is a research direction that can be pursued. One can also extend the results of this study to find optimal locations of new vaccination sites. This can be tricky because the optimization problem will become continuous and hence, other numerical techniques must be employed.

Although the method is intended for COVID-19 vaccinations, the method is general enough that it can be applied to formulating vaccination strategies of other diseases. For example, if the vaccination is only for school-age children, (this is applicable for diseases like soil-transmitted helminths and schistosomiasis), then the locations can be restricted to just the elementary schools.

We hope that this study can help stakeholders in planning strategies to end the COVID-19 pandemic, which has crippled the world economy and has affected the lives of millions of people worldwide.

## Data Availability

The data used in the numerical method section can be found in the Supplementary File at the end of the manuscript file.

## 6 DECLARATIONS

### Availability of data and material

The data used in the numerical method section can be found in the Supplementary File.

### Code availability

The code and the tutorial for the optimization implementation can be found in the following GitHub repository: https://github.com/kurtizak/Covid-Site-Optimization

### Authors’ contributions

- K.I.M. Cabanilla and E.A.T. Enriquez conceived and designed the experiments, performed the experiments, analyzed the data, performed the computation work, prepared figures and/or tables, authored or reviewed drafts of the paper, and approved the final draft.
- R. Mendoza and V.M.P. Mendoza conceived and designed the experiments, analyzed the data, authored or reviewed drafts of the paper, and approved the final draft.

## Supplementary File for the paper

San Juan is a municipality in the province of Batangas, located in the Luzon island of the Philippines. It consists of 42 barangays. A map of San Juan, Batangas is shown in Figure 1.

**Figure 1.**
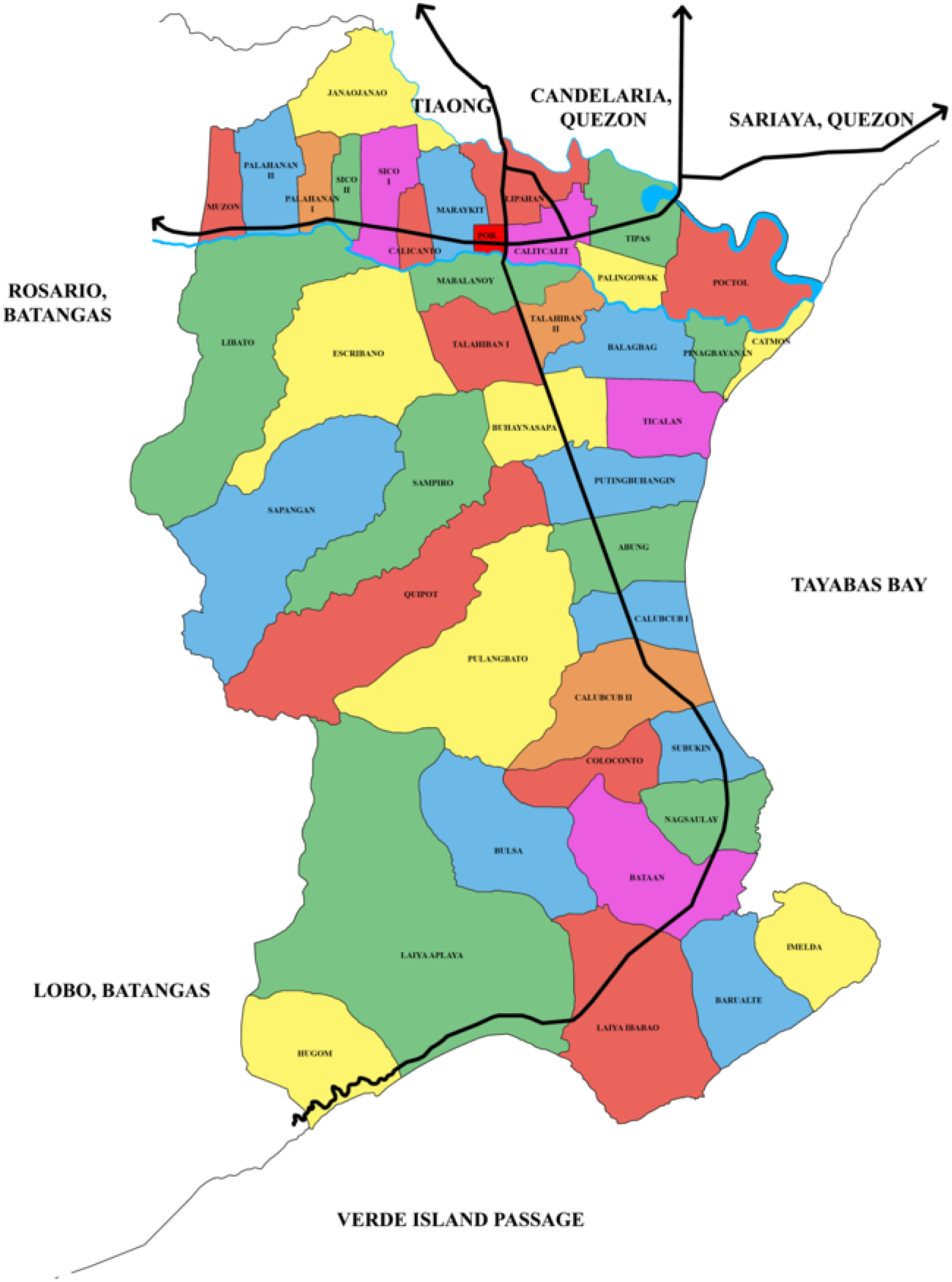
Map showing the barangays of San Juan, Batangas, the Philippines. The image was made by Marvin Sinag in 2019 and was obtained from https://commons.wikimedia.org/wiki/File:Ph_map_of_san_juan_batangas.png

The population data were collected from the website of the Philippine Department of Health (https://doh.gov.ph/publications). The number of COVID-19 confirmed cases per barangay in San Juan, Batangas was obtained from the Facebook page of the San Juan local government unit (https://www.facebook.com/lgusanjuanbatangas).

The list of hospitals and schools was manually generated from the directories of the Philippine Department of Health (https://nhfr.doh.gov.ph/rfacilities2list.php?pageno=1&t=rfacilities2&recperpage=ALL), Philippine Commission on Higher Education (https://ched.gov.ph/list-higher-education-institutions/), and the Philippine Department of Education (https://www.deped.gov.ph/k-to-12/senior-high-school/list-of-senior-high-schools/). The coordinates of the barangay halls and vaccination sites were collected with the help of the OSM package in Python and Google maps

**Table I:** All the barangays in San Juan, Batangas with their projected population in 2021, number of confirmed COVID-19 cases, and coordinates.

|  |  |  |  |  |
| --- | --- | --- | --- | --- |
| **NAME** | **POPULATION** | **NO. OF CASES** | **LATITUDE** | **LONGITUDE** |
| Abung | 2,444 | 14 | 13.7685 | 121.4156 |
| Balagbag | 2,929 | 7 | 13.8032 | 121.4165 |
| Barualte | 1,933 | 2 | 13.6874 | 121.4389 |
| Bataan | 2,204 | 3 | 13.6964 | 121.4333 |
| Buhay Na Sapa | 5,909 | 26 | 13.7905 | 121.4081 |
| Bulsa | 2,492 | 3 | 13.7027 | 121.4003 |
| Calicanto | 2,257 | 13 | 13.8258 | 121.3821 |
| Calitcalit | 5,255 | 59 | 13.8268 | 121.4111 |
| Calubcub I | 2,545 | 13 | 13.7534 | 121.4211 |
| Calubcub II | 4,071 | 9 | 13.7421 | 121.4262 |
| Catmon | 1,636 | 0 | 13.8066 | 121.4503 |
| Coloconto | 874 | 8 | 13.7228 | 121.4192 |
| Escribano | 3,755 | 8 | 13.8122 | 121.3644 |
| Hugom | 1,820 | 7 | 13.6631 | 121.3731 |
| Imelda (Tubog) | 1,106 | 3 | 13.6900 | 121.4510 |
| Janaojanao | 1,743 | 1 | 13.8539 | 121.3677 |
| Laiya-Aplaya | 6,927 | 14 | 13.6762 | 121.3876 |
| Laiya-Ibabao | 5,195 | 11 | 13.6807 | 121.4161 |
| Libato | 4,737 | 10 | 13.8156 | 121.3511 |
| Lipahan | 5,052 | 42 | 13.8339 | 121.4056 |
| Mabalanoy | 4,423 | 38 | 13.8183 | 121.3975 |
| Maraykit | 4,123 | 42 | 13.8258 | 121.3872 |
| Muzon | 1,775 | 11 | 13.8281 | 121.3409 |
| Nagsaulay | 3,004 | 21 | 13.7117 | 121.4404 |
| Palahanan I | 970 | 2 | 13.8297 | 121.3608 |
| Palahanan II | 3,730 | 13 | 13.8303 | 121.3523 |
| Palingowak | 1,883 | 7 | 13.8217 | 121.4136 |
| Pinagbayanan | 1,739 | 3 | 13.8008 | 121.4367 |
| Poblacion | 3,785 | 76 | 13.8276 | 121.3947 |
| Poctol | 2,939 | 16 | 13.8205 | 121.4408 |
| Pulangbato | 3,192 | 1 | 13.7472 | 121.3967 |
| Putingbuhangin | 2,873 | 22 | 13.7819 | 121.4231 |
| Quipot | 3,839 | 6 | 13.7709 | 121.3860 |
| Sampiro | 3,635 | 7 | 13.7866 | 121.3924 |
| Sapangan | 3,391 | 15 | 13.7840 | 121.3582 |
| Sico I | 2,280 | 19 | 13.8282 | 121.3732 |
| Sico II | 1,269 | 8 | 13.8295 | 121.3632 |
| Subukin | 1,886 | 2 | 13.7298 | 121.4380 |
| Talahiban I | 2,588 | 3 | 13.8089 | 121.3931 |
| Talahiban II | 1,501 | 3 | 13.8095 | 121.4039 |
| Ticalan | 2,111 | 6 | 13.7924 | 121.4268 |
| Tipaz | 3,432 | 5 | 13.8317 | 121.4283 |
| **TOTAL** | 125,252 | 579 |  |  |

**Table II:**
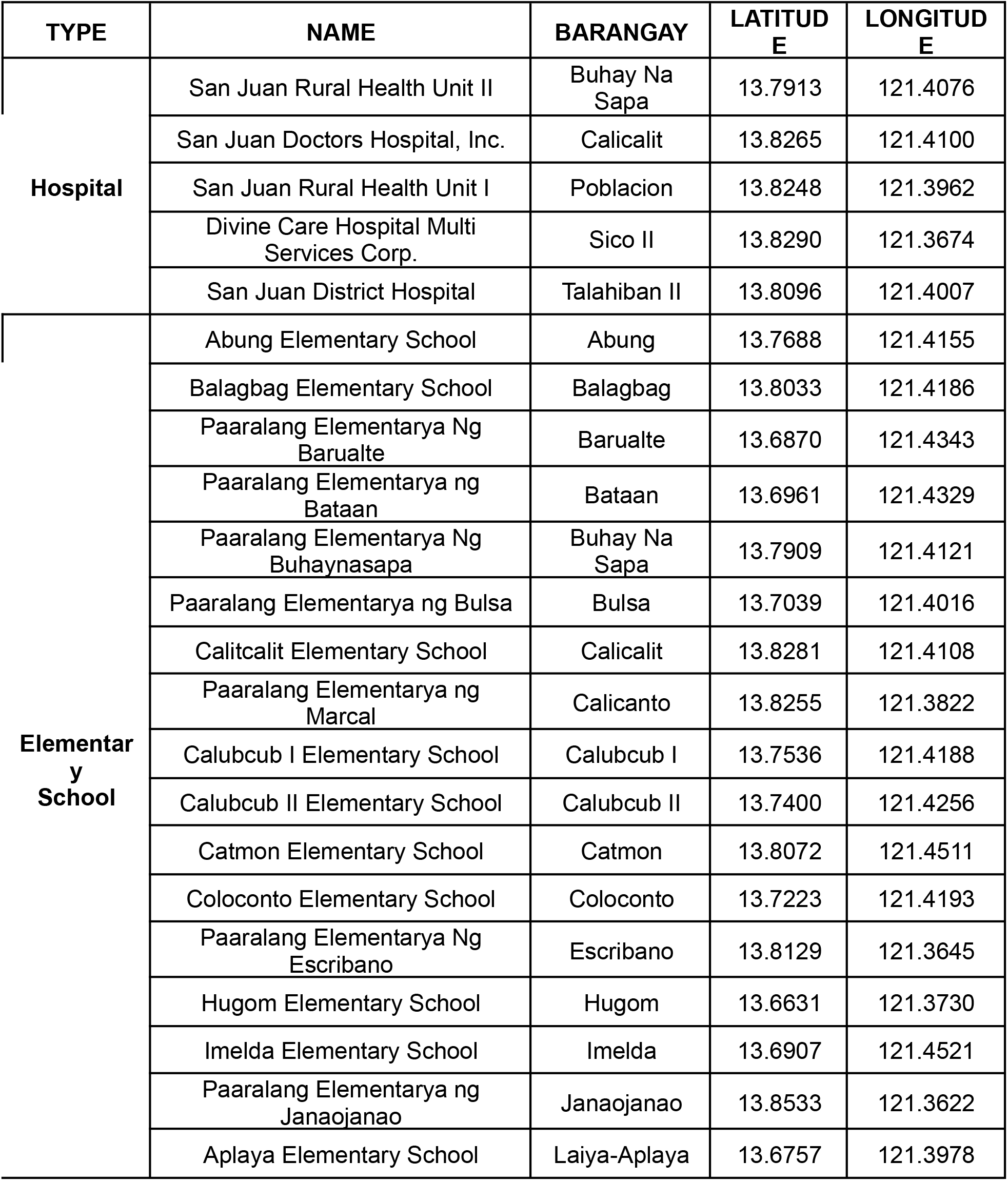
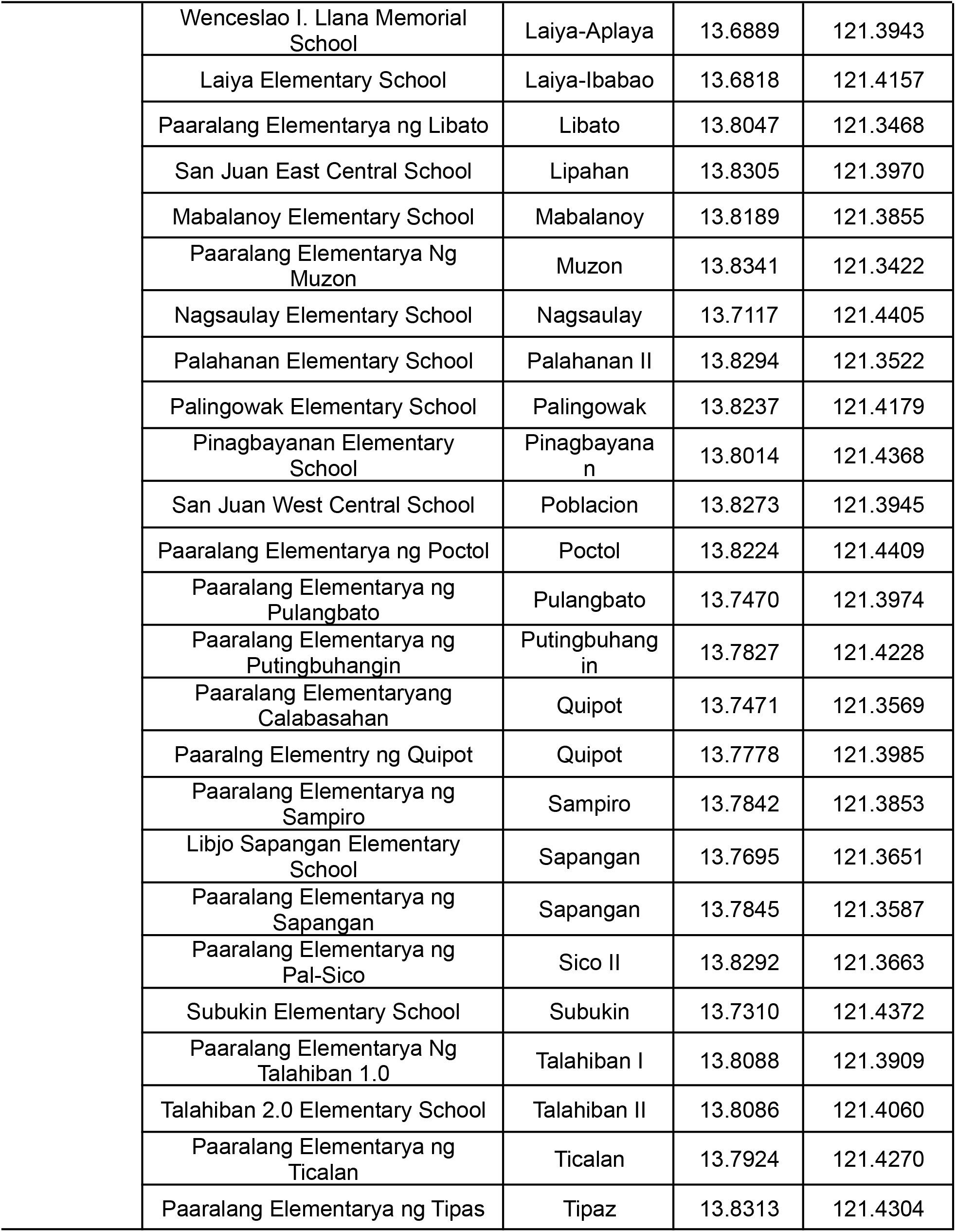
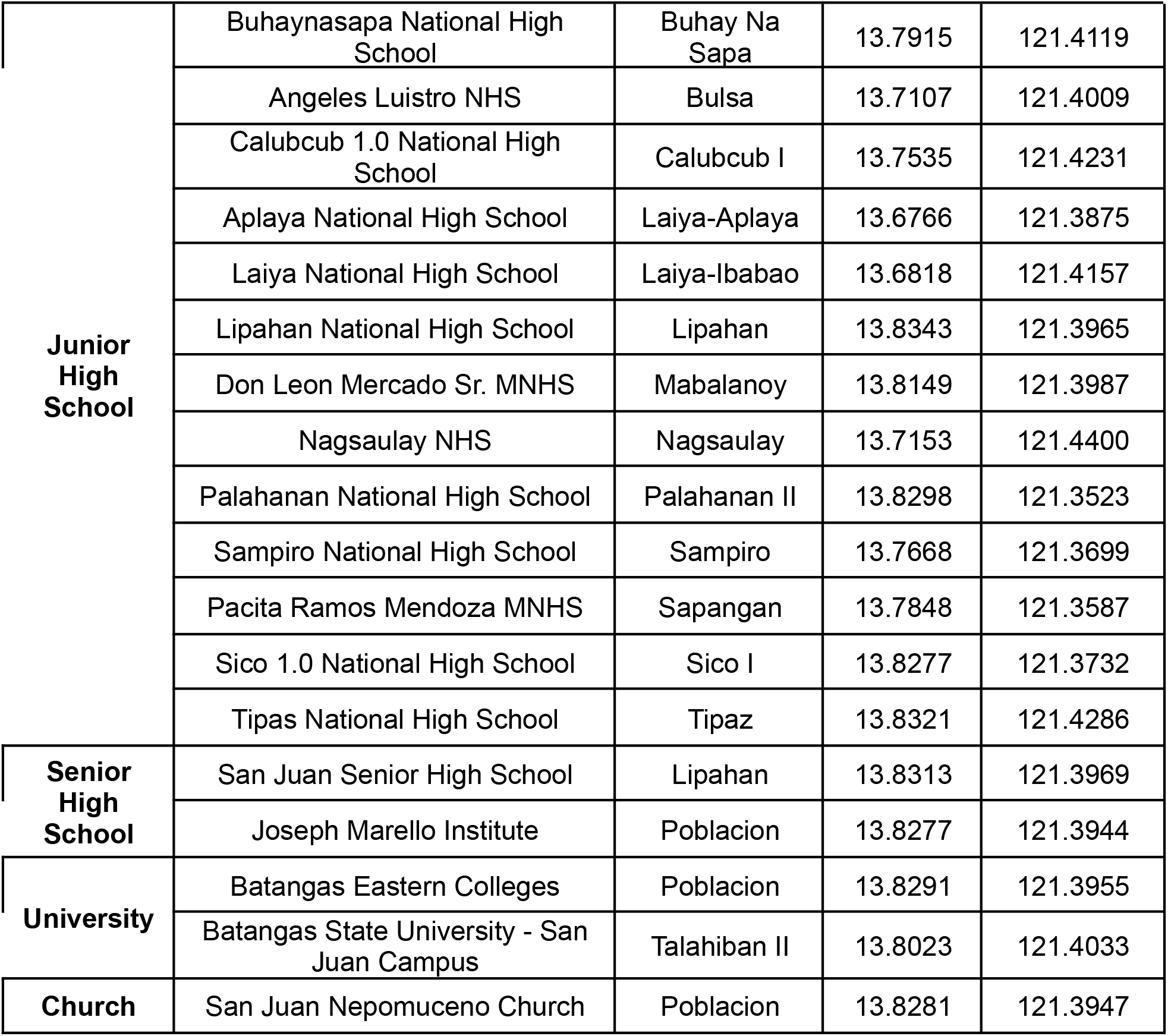
Names of the vaccination sites in San Juan, Batangas with their corresponding site type, barangay location, and coordinates.

